# A broad-based probe-free qPCR assay for detection and discrimination of three human herpes viruses

**DOI:** 10.1101/2020.10.01.20205427

**Authors:** Anshu Gupta, Shelley M. Lawrence, Stephanie I. Fraley

## Abstract

Human cytomegalovirus (HCMV) and herpes simplex viruses (HSV) 1 and 2 are double stranded DNA viruses that establish lifetime latency in a host upon initial infection. Primary infection or reactivation of latent virus during pregnancy can transmit the virus *in utero* or during natural childbirth to the fetus. The majority of these infections are asymptomatic at birth but may present later with potentially lethal disseminated infection or meningitis (HSV), or long-term neurodevelopmental sequelae including sensorineural hearing loss or neurodevelopmental impairments (HCMV). Unfortunately, early signs and symptoms of disseminated viral infections may be misdiagnosed as bacterial sepsis. Therefore, immediate testing for viral etiologies may not be ordered or even considered by skilled clinicians. In asymptomatic HCMV infections, early detection is necessary to monitor for and treat future neurologic sequelae. In acutely ill-appearing infants, specific detection of viruses against other disease-causing agents is vital to inform correct patient management, including early administration of the correct antimicrobial(s). An ideal test should be rapid, inexpensive, require low sample volumes, and demonstrate efficacy in multiple tissue matrices to aid in timely clinical decision-making for neonatal infections. This work discusses the development of a rapid probe-free qPCR assay for HSV and HCMV that enables early and specific detection of these viruses in neonates. The assay’s probe free chemistry would allow easier extension to a broad-based multiplexed pathogenic panel as compared to assays utilizing sequence-specific probes or nested PCR.

## Introduction

Human cytomegalovirus is the most common cause of congenital viral infection in humans and the leading non-genetic etiology of neurodevelopmental sequelae in children, including intellectual disability, cerebral palsy, seizures, visual defects and sensorineural hearing loss (1–8). HCMV seroprevalence among women of reproductive age is high, ranging from 45-100% worldwide (9). In the United States, an estimated 58.3% of pregnant women harbor the virus (10), with an additional 27,000 new CMV infections reported in pregnant women each year (11). The infection is usually asymptomatic but active viral replication in pregnant women, resulting from new (primary) infections or reinfection/reactivation of latent virus (non-primary infection) can lead to congenital CMV infection or cCMV in the fetus. While the risk for cCMV from latent primary infections is <3%, it can range from 30-70% for primary infection or viral reactivation (9). Therefore, one in every 200 babies born in the US will be infected with cCMV according to CDC data (https://www.cdc.gov/cmv/clinical/congenital-cmv.html), or an estimated 30,000 cases annually.

Although the majority (90%) of neonates with cCMV infection are asymptomatic at birth, an estimated 1 in 10 of well-appearing cCMV-positive infants will develop late onset health conditions, including sensorineural hearing loss (SNHL), months to years after their initial infection (12). The remaining 10% of infants infected with cCMV *in utero* will present with clinical symptoms at birth (12) and nearly half will develop significant long term neurodevelopmental challenges (from CDC, https://www.cdc.gov/cmv/clinical/congenital-cmv.html). Most cases (>50%) of cCMV resulting in SNHL are progressive in nature and may be missed by standard, routine hearing screens at the time of birth (8). As symptoms of progressive hearing loss and/or clinical presentation of neurologic disabilities are not specific to HCMV, clinicians may order multiple diagnostic tests and/or ineffective therapies in the quest to diagnose other causes, which can substantially increase medical costs, while failing to identify the actual cause (1). cCMV infections also need to be diagnosed within the first three weeks of life, after which, congenital infections cannot be distinguished from postnatally acquired infections, which are usually not associated with neurologic sequelae (13).

Rapid, inexpensive, specific and early detection of cCMV is thus required to manage disease progression and inform early intervention. First, earlier CMV diagnosis can help initiate timely therapy and prevent/decrease the development of sequelae, particularly in case of asymptomatic and late onset conditions. Second, cCMV screening would help to monitor future hearing loss and provide timely intervention, which has been known to improve receptive and expressive language skills (1). Third, early diagnosis would incur significant cost savings by avoiding extraneous diagnostic tests or therapies in the quest to find the actual source of CMV caused neurodevelopmental impairments (1). Fourth, a precise and rapid CMV diagnostic test would also aid in antiviral clinical trials by identifying the neonates that may benefit from this therapy.

Like HCMV, herpes simplex viruses (HSV) are types of herpes viruses that are known to infect humans. There are two types of herpes simplex viruses: (a) HSV1 that has been traditionally associated with oral, labial, and facial lesions and (b) HSV2 that is the common etiology of genital herpes. However, recent studies indicate HSV1, a more virulent strain than HSV2, is rapidly rising as a primary etiology of genital herpes, leading to 20-50% of current cases (14, 15). HSV, like HCMV, establishes latency in the host with a particular predilection for neurons of dorsal root ganglia and autonomic nervous system (16).

Seroprevalence of HSV infections in women of ages 15-49 in the US is 50.9% for HSV1 and 15.9% for HSV2 based on CDC (https://www.cdc.gov/nchs/products/databriefs/db304.htm). Two in three women are either asymptomatic or show ambiguous symptoms which makes it difficult to diagnose and treat the infection (17, 18). HSV can be transmitted from a pregnant woman to the neonate trans-placentally *in utero* in 5% cases, through contact with infected genital secretions peripartum in 85% cases or postnatally in 10% infections (19). Like CMV, transmission risk is higher for primary infections (57%) as compared to 25% for non-primary infections and 2% for recurring infections (18). Unlike HCMV, however, HSV transmission does not occur via ingestion of breast milk (20).

Neonatal HSV infection (nHSV) is less common than cCMV, with occurrence in every 1 in 3200 births annually in the US, or around 1500 nHSV cases every year (18, 20). nHSV may present as skin-eye-mouth disease (SEM; 45%) such as keratitis and conjunctivitis, disseminated disease (25%) or neurologic disease (30%), including meningitis and encephalitis (20, 21). Without therapy, the mortality rate associated with disseminated disease is 85% and 50% for central nervous system (CNS) disease, which is greatly reduced to 31% and 6%, respectively, with antiviral therapy (22). This underscores the need for early diagnosis and timely therapy for nHSV. HSV is also the major cause of viral sepsis and has many common non-specific symptoms with other sepsis causing pathogens. It may, therefore, be difficult to differentiate HSV from more common pathogens that cause neonatal sepsis, particularly in the absence of skin lesions (23). The gold standard in sepsis detection is blood culture (24), which cannot detect HSV and other viral agents.

An early and rapid test for HSV is thus necessary for the following reasons. First, it will allow early diagnosis and intervention of otherwise asymptomatic infections in neonates. Second, it can help specific detection and correct treatment for neonatal sepsis in case of non-specific symptoms, which may otherwise be misdiagnosed or left undiagnosed. Third, it can be used to rapidly screen laboring women for active HSV infection that will inform the safest mode of delivery. In case of infection, cesarean delivery could be ordered to reduce peripartum transmission risk through the birth canal. The newborn can also be rapidly identified as at risk for HSV infection and appropriate postnatal observation and clinical management initiated.

Therefore, the aim of this work was to develop a probe-free universal real time PCR based assay to detect HCMV and HSV1 and 2 through high resolution melt (HRM) technology. The current gold standard for HCMV and HSV1 and 2 diagnosis is viral culture (25), which does not satisfy the requirements for a rapid test for early neonatal detection as discussed above. These tests have slower turn-around times that makes them unsuitable as a screening tool, have decreased sensitivity (26), higher costs than molecular tests and require larger sample volumes. Multiplexed molecular tests are also used in some laboratories. They usually utilize universal primers or multiple pathogenic primers added together in the same reaction, along with either nested PCR (same external but different species-specific internal primers) or sequence specific fluorescent probes to differentiate each species (specificity) (27). This limits the number of pathogens that can be detected in a single reaction chemistry due to a cap on the maximum number of different oligonucleotides (primers or probes) that can be used effectively in the same reaction. Some assays split multiple primers/probes across different reactions, leading to a larger sample volume requirement to maintain assay sensitivity.

This work hypothesizes the use of probe-free assays with universal primer chemistry and specificity based on high-resolution melt technology to alleviate the above limitations on broad based pathogen detection. Instead of sequence specific probes, species level identification is accomplished by high resolution melt signature that uses non-specific fluorescent intercalating dyes. The use of probe free assays would also allow extension of the assay to include more pathogens without redesigning new probes for each new species. Here, a universal probe free quantitative PCR (qPCR) assay was designed to detect HCMV, HSV1 and HSV2 utilizing high-resolution melt technology. The work involves primer design from viral sequences, qPCR assay optimization for the primers and its analytical characterization. The assay would help in improving neonatal disease management through early detection and timely intervention, specifically in cases of asymptomatic infections. The inherent rapidity of the qPCR test as compared to viral culture and its higher sensitivity and cost effectiveness also allow it to be used as a screening tool for maternal viral infections during pregnancy or for universal newborn screening for these viruses.

## Materials and Methods

### Design of primers

To find universal primers for the three human herpesviruses, 6 gene families whose products are evolutionarily conserved in these viruses were considered: (a)capsid, (b)tegument and cytoplasmic egress, (c)envelope, (d)regulation, (e)DNA replication, recombination and metabolism, and (f)capsid assembly, DNA encapsidation and nuclear egress (28). Each gene from the six selected gene families was manually aligned for the three viruses (HCMV:NC_006273.2, HSV1: NC_001806.2 and HSV2: NC_001798.2) using the online Benchling alignment tool (http://www.benchling.com) to find all possible primers conserved in the three species. The primers were analyzed for homo-dimerization, hetero-dimerization and hairpin formation *in silico* using online tools: IDT oligo analyzer, Sigma Aldrich OligoEvaluator and PREMIER Biosoft NetPrimer, to obtain 51 primer pairs. To our knowledge, only one of these primers is already found in literature (29) (Table 1). Barring one primer pair from the Glycoprotein B gene, all the primers are from the highly conserved DNA Polymerase gene of the viruses (27). The primers range in length from 18-30 bases.

**Table 1:**
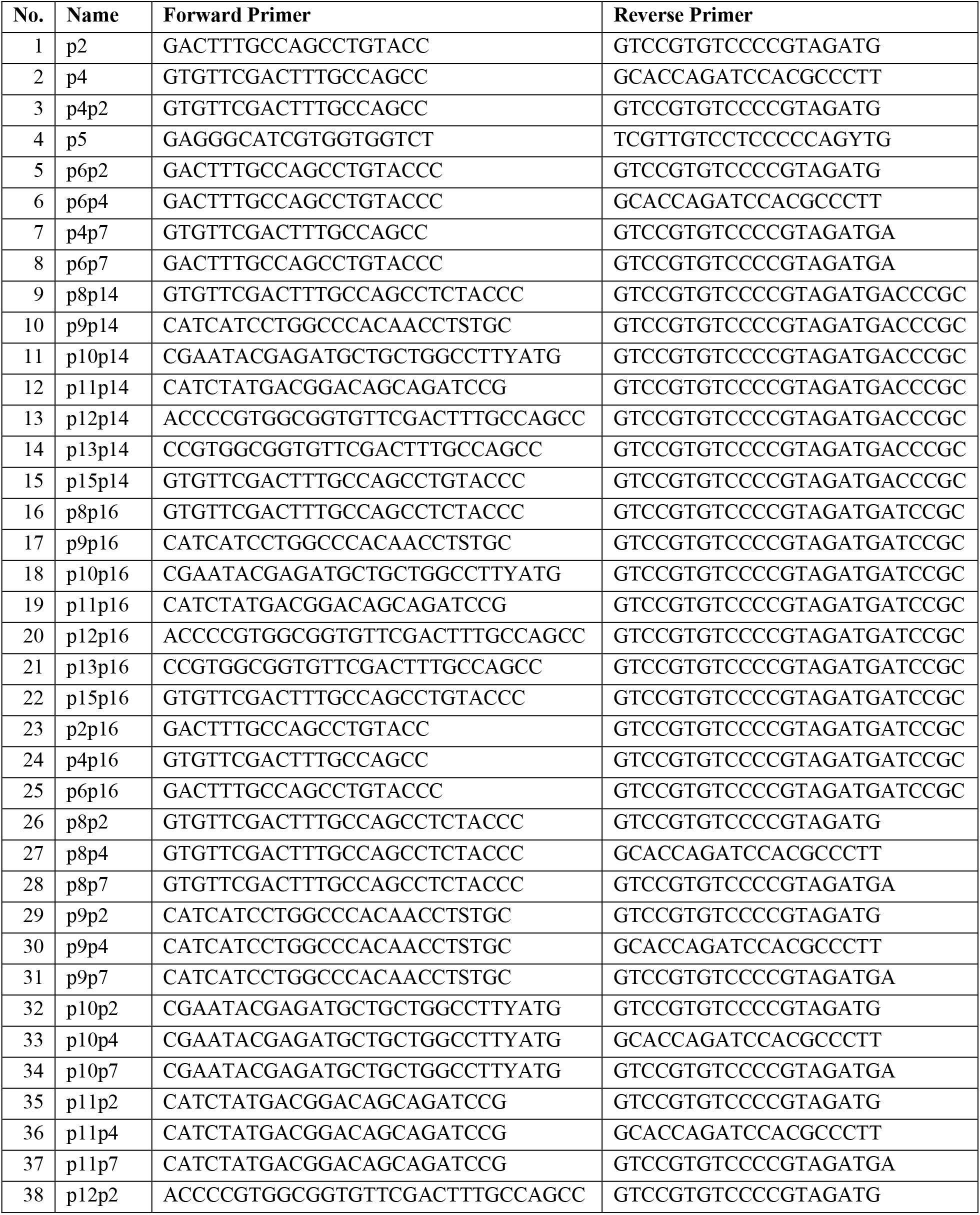
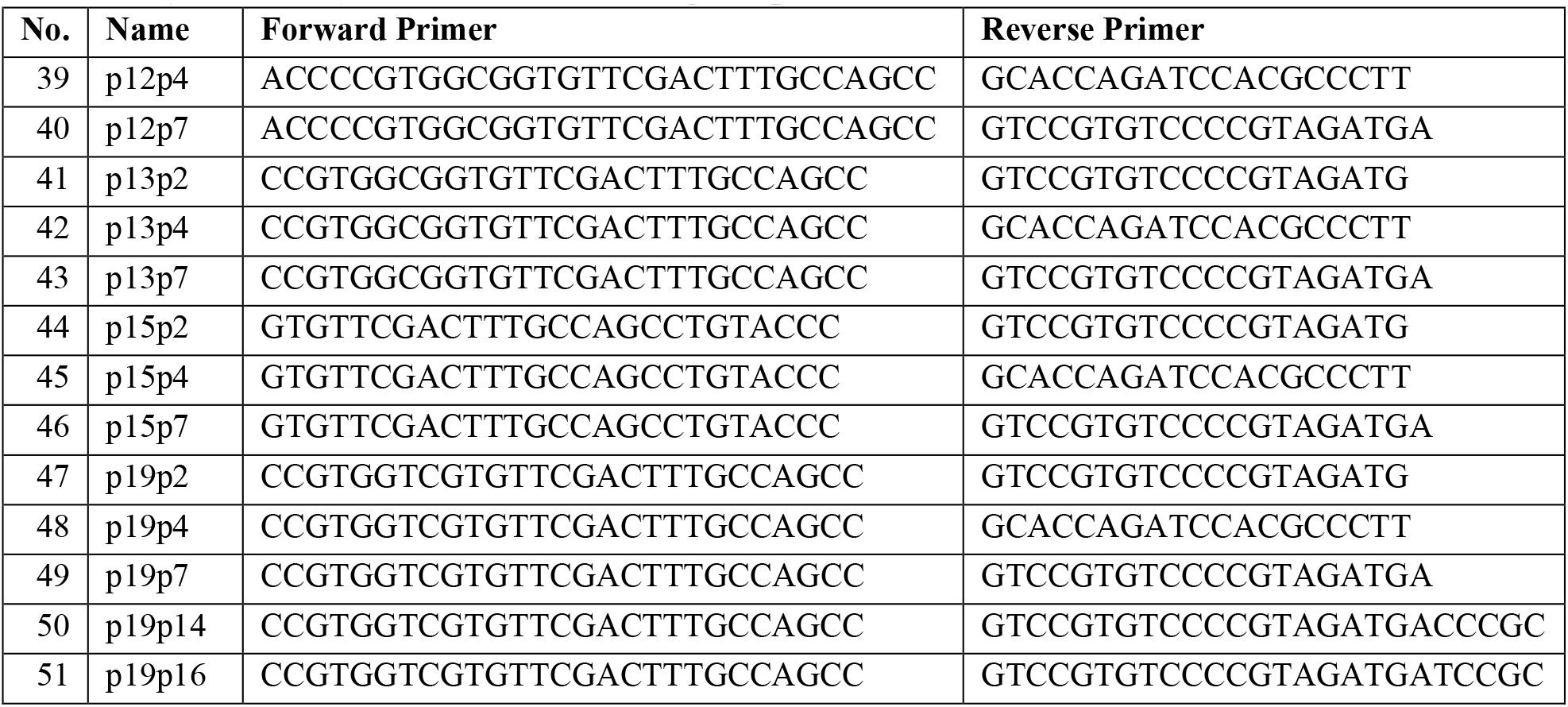
List of custom designed primers

**Table 2:** Characteristics of the viral assay for HCMV, HSV1 and HSV2 amplification

|  | Linear Dynamic Range | LOQ (copies/reaction) | R<sup>2</sup> | Efficiency (%) | Slope | Y intercept | Mean Cq at LOQ |
| --- | --- | --- | --- | --- | --- | --- | --- |
| HCMV | 10<sup>7</sup>- 10<sup>3</sup> | 10<sup>3</sup> | 0.987 | 76.92 | -4.04 | 41.45 | 30 |
| HSV1 | 10<sup>7</sup>- 10<sup>4</sup> | 10<sup>4</sup> | 0.991 | 85.022 | -3.74 | 43.25 | 28 |
| HSV2 | 10<sup>6</sup>- 10<sup>4</sup> | 10<sup>4</sup> | 0.905 | 58.089 | -5.03 | 50.50 | 30 |

### Human and viral DNA

Human genomic DNA for specificity experiments was extracted from frozen human cord blood samples obtained from Rady Children’s Hospital. The frozen blood was thawed and DNA extracted in multiple aliquots using Promega Wizard Genomic DNA Purification kit using the vendor recommended protocol for whole blood to obtain an average of 160µg/mL DNA in PCR purified water. Extracted viral DNA was obtained from ATCC with the following product details: HCMV strain AD-169-ATCC VR-538D, HSV1 strain KOS-ATCC VR-1493D and HSV2 strain G-ATCC VR-734.

### Testing of primer specificity against human DNA

All 51 primer pairs were tested against human DNA amplification. Each primer set was tested in four reactions, two with no template (no template controls or NTC) and two with extracted human genomic DNA as the template. All reaction volumes were 15µL with 1x NEB Q5 buffer, 2.5x EvaGreen, 0.2mM dNTPs, 0.02U/µL of NEB Q5 Polymerase and 0.5µM each of forward and reverse primers. Human DNA was used at a final reaction concentration of 3.2ng/µL. Thermocycling was performed in Thermo Fisher QuantStudio Real-Time PCR System at the following conditions: 1.5 minutes of initial denaturation at 98°C, 55x cycles of 10 seconds denaturation at 98°C, 30 seconds annealing at varying temperatures and 30 seconds elongation at 72°C, followed by a 5-minute final extension at 72°C. Post the amplification cycles, the reactions were heated to 95°C followed by a melting curve temperature gradient from 60°C to 99.9°C. The annealing temperatures were estimated from NEB Tm calculator (http://www.tmcalculator.neb.com) to be 69°C for primer sets 1-8, 72°C for primer sets 9-22 and 47-51 and 70°C for primer sets 23-46.

Amplification and melt curves for each primer pair were analyzed and gel electrophoreses performed to find primers that do not amplify human DNA. The selected primer pairs were then re-subjected to PCR in 12 reactions-2 as NTCs and 10 with human DNA as the template. Primer sets that did not amplify human DNA were selected for further testing.

### Real time PCR assay development for herpes viruses

Selected primer sets that did not amplify human DNA were tested for HCMV (1.53×10^5^ genome copies/reaction) and HSV1 (5.23×10^5^ genome copies/reaction) amplification in duplicates with 2 separate reactions as NTC. The thermocycling conditions for amplification and melting are the same as followed previously. Analysis was performed on their melt and amplification curves and gel electrophoresis. The primer set showing specific amplification of the two viruses were chosen and retested in qPCR in replicates of 6 for each virus. HSV2 testing was conducted after the designed and selected primer assay was optimized, since HSV2 genome is similar to HSV1.

### Optimization of assay with the selected primer pair

The selected primer assay was optimized for various parameters, including the DNA Polymerase, primer annealing temperature, primer concentration, amplification thermocycling temperature ramp rates and extension times. All the optimization reactions were conducted with HCMV and HSV1 DNA.

For comparing the DNA Polymerase enzymes, 10X serial dilutions of HCMV and HSV1 ranging from 10^5^ to 10^2^ genome copies/reaction were amplified in triplicates each with NEB Phusion Hot Start DNA Polymerase and NEB Q5 High Fidelity DNA Polymerase. The reaction conditions were as previously stated with an annealing temperature of 70°C for both Polymerases.

For annealing temperature optimization, 10X serial dilutions of HCMV and HSV1 ranging from 10^4^ to 10^2^ genome copies/reaction each were amplified in duplicates using the Polymerase selected from the previous step. The experiment was carried out in Bio-Rad CFX96 qPCR instrument for 6 different temperatures ranging from 71.7°C to 67.4°C as follows: 71.7°C, 71.2°C, 70.2°C, 69.0°C, 68.0°C and 67.4°C.

Primer concentration experiments were performed with HSV1 by designing a matrix of different primer concentrations ranging from 0.05 to 0.5µM/reaction for forward and reverse primers. Both asymmetric as well as symmetric primer concentrations were tested. Ramp rates and extension time were also optimized for maximum sensitivity.

### Analytical characterization of the assay to assess the limit of quantification, dynamic range and reproducibility

Using the selected primer pair, standard curves were created for eight 10X serial dilutions in triplicates for HCMV and HSV1: 8.45×10^7^ to 8.45×10° genome copies/reaction for HCMV and 5.23×10^7^ to 5.23×10° genome copies/reaction for HSV1. For HSV2, seven 10x serial dilutions were used, ranging from 3.45×10^6^ to 3.45×10° genome copies/reaction. The qPCR standard curves were used to find the linear dynamic range, limit of quantification (LOQ), Ct corresponding to LOQ and the reaction efficiency. The reproducibility of the reaction was found by inter and intra reaction variability through repeat experiments (triplicates for HCMV and HSV2 and duplicates for HSV1).

## Results

### Design of universal primers

Of the 51 primer pairs designed, only eight were found to be experimentally specific against human genomic DNA and did not amplify it in initial as well as repeat experiments. However, four of these (p12p2, p12p7, p13p2 and p13p14) formed multiple dimers and/or extraneous PCR products, as observed from the multiple melt curve peaks and multiple bands in gel electrophoresis (Figure 1).

**Figure 1:**
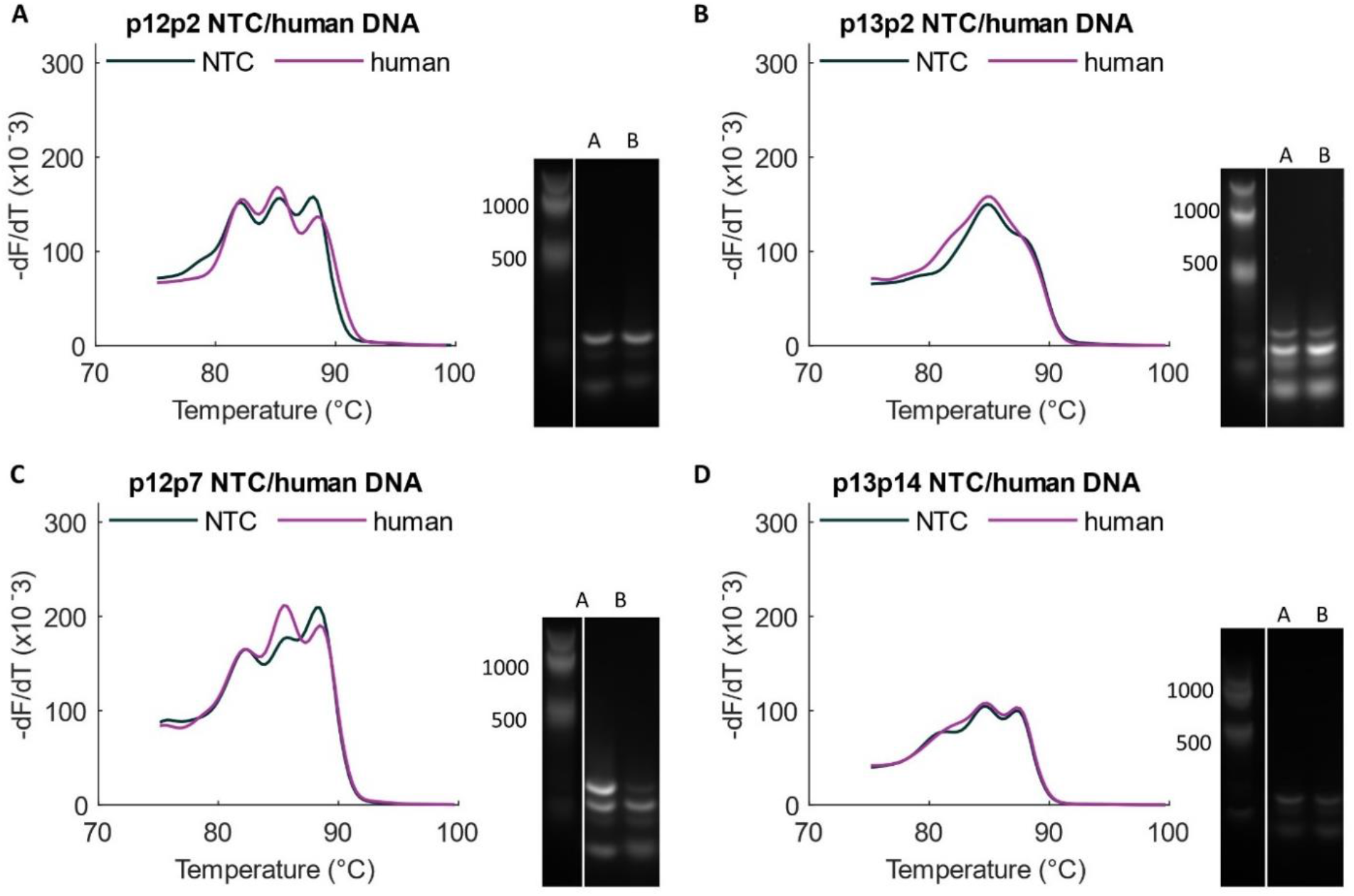
Primer dimer formation by custom primers. (A to D) Melt curves and gels showing formation of multiple primer dimers by p12p2, p13p2, p12p7 and p13p14, respectively. Columns A and B in the gels represent NTC and Human DNA as template, respectively.

The remaining four of the eight selected primer pairs were tested with HCMV and HSV1. With three of these (p11p2, p11p7 and p11p16), the forward primer, p11, bound to HSV1 at more than one position giving multiple products, as observed by multiple product peaks in the melt curves and bands on gel electrophoresis.

The remaining eighth primer set, labelled p19p4, showed specific amplification of all three viruses without amplification of human DNA (Figure 3). The probe-free specificity can be observed as distinct melting signatures for the three viral amplicons with different average melting temperatures as follows: 94.33±0.32°C for HCMV, 95.70±0.28°C for HSV1 and 96.00±0.14°C for HSV2.

**Figure 2:**
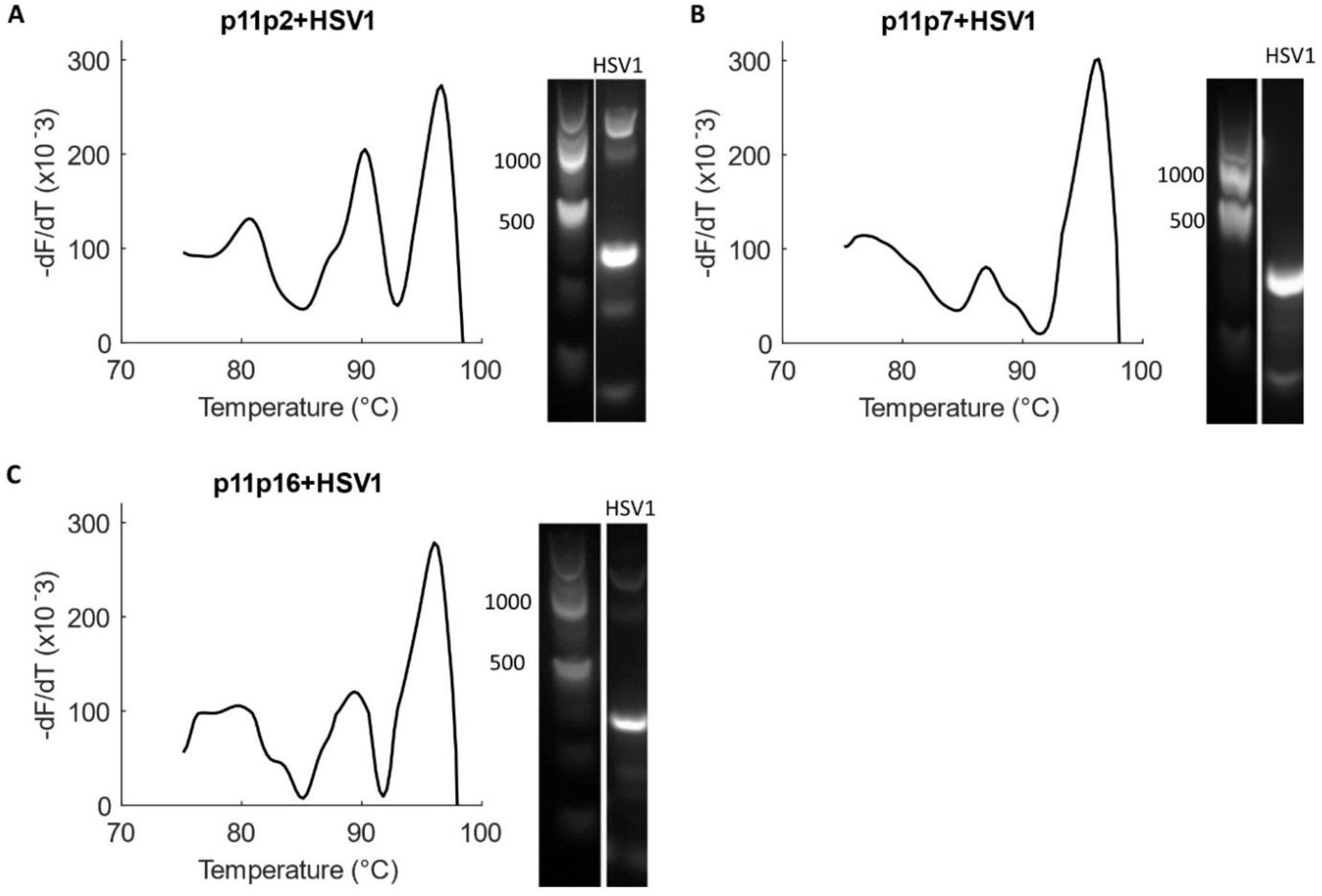
Primer p11 forms multiple products with HSV1. (A to C) p11p2, p11p7, p11p16 form multiple products with HSV1 as can be seen from their multiple melt peaks as well as bands in gel electrophoresis. Part of the forward primer, p11, was found to bind at multiple positions to HSV1 giving more than one product. The actual product in all cases was 829 bp long whereas both longer and shorter products are also observed.

**Figure 3:**
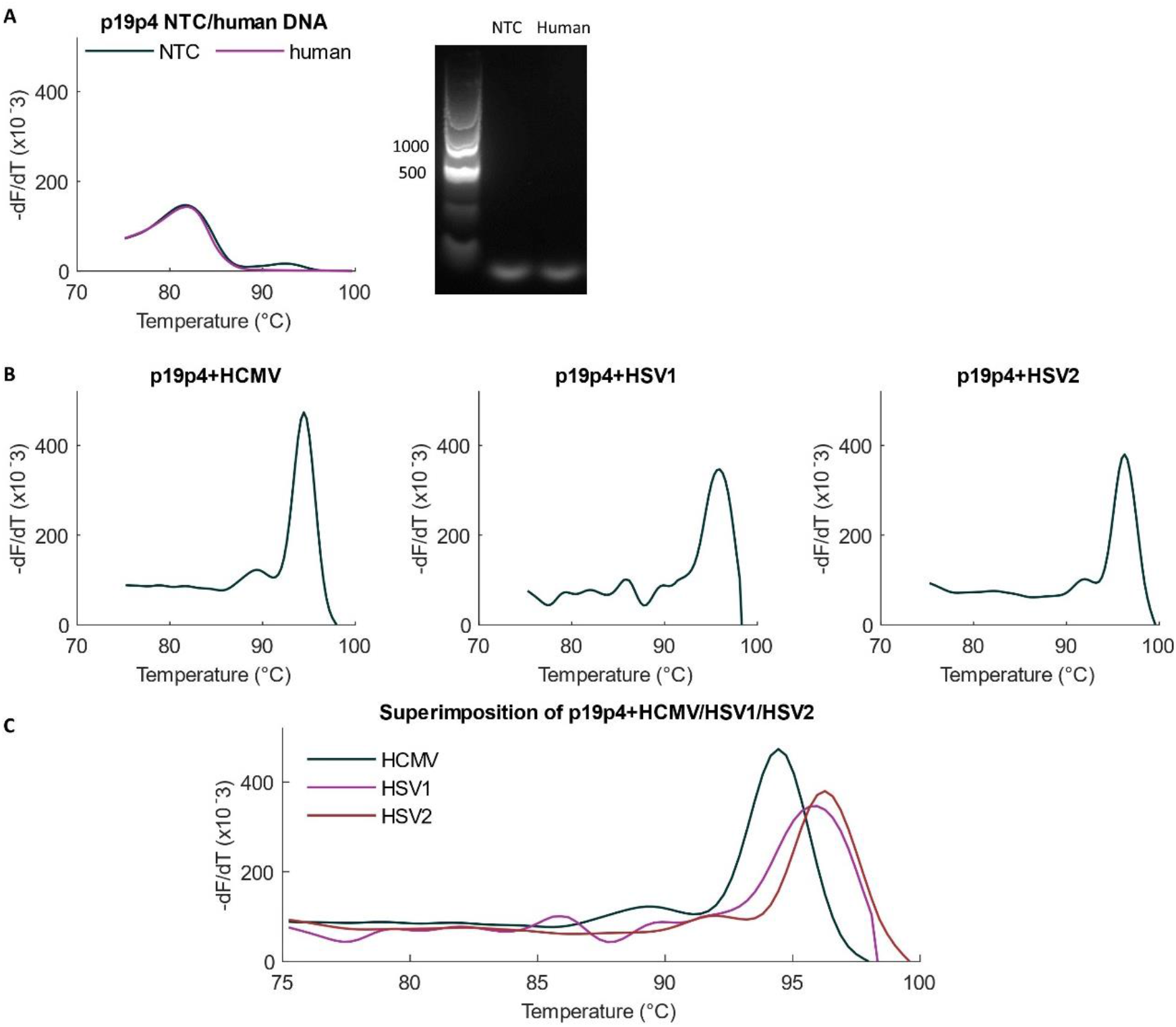
Amplification with primer pair p19p4. (A) The primer pair named p19p4 is specific against human DNA amplification as shown by the alignment for melt curves in NTC and human DNA as template. There was no product melt peak observed with human DNA although the primer forms a primer dimer. (B) Melt curves for each virus with the primer pair p19p4. (C) The melt curves for the three viruses are superimposed together to show their distinct signatures.

### Optimization of the DNA Polymerase chemistry of the assay

Although initial amplification experiments were performed with Q5 DNA Polymerase, we also wished to test the performance of the assay with High Fidelity NEB Phusion Hot Start DNA Polymerase. With Phusion, the lowest concentration at which product melt peaks were observed, or the limit of detection (LOD), was in the order of 10^4^ viral genome copies/reaction for both HCMV and HSV1-1.53×10^4^ copies/reaction for HCMV and 5.23×10^4^ copies/reaction for HSV1. LOD for Q5 was an order lower (better) at 10^3^ viral genome copies/reaction for both viruses (HCMV - 1.53×10^3^ copies/reaction, Figure 4 and HSV1 - 5.23×10^3^ copies/reaction, Figure 5).

**Figure 4:**
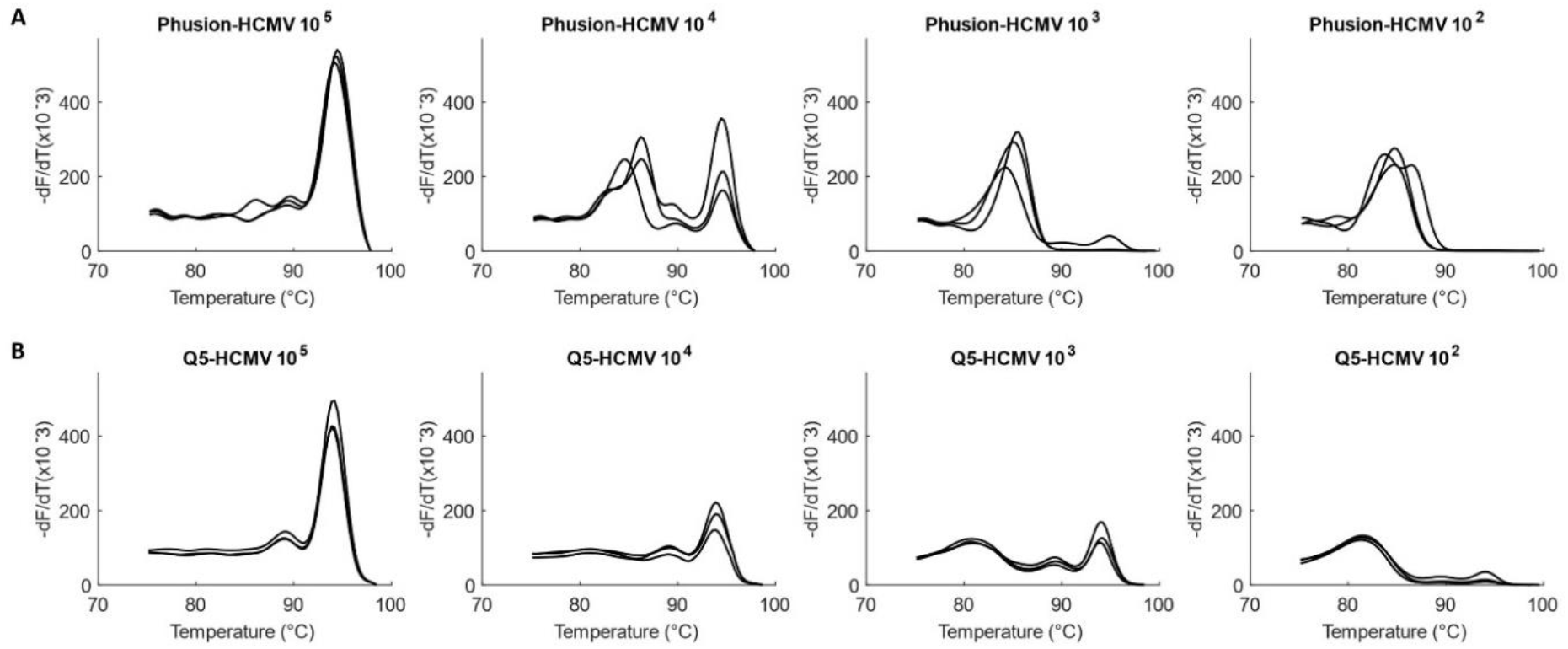
Comparison of Phusion vs Q5 DNA Polymerases for amplification of HCMV. Amplification of serially diluted HCMV (1.53×10^5^ to 1.53×10^2^ copies/reaction) in triplicates with (A) Phusion-LOD of 1.53×10^4^ copies and with (B) Q5-LOD of 1.53×10^3^ copies.

**Figure 5:**
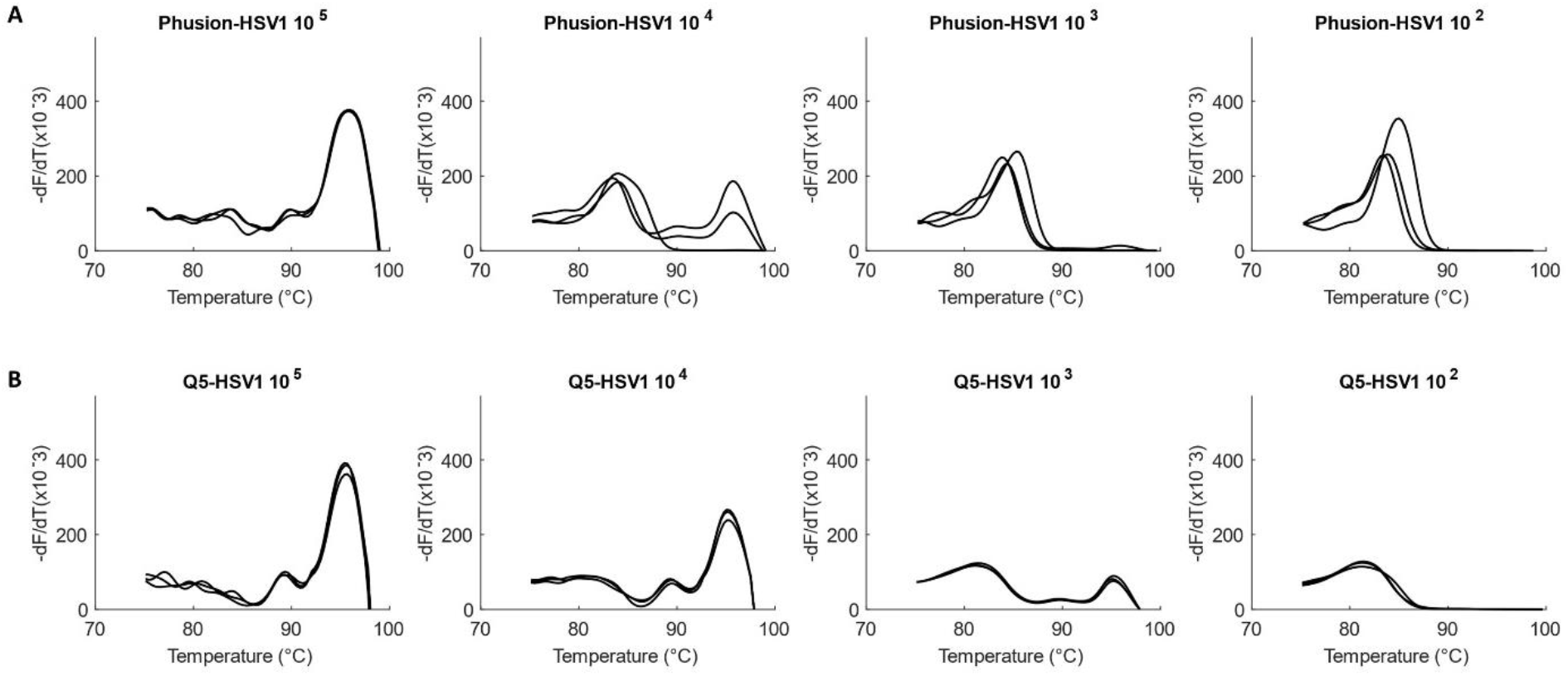
Comparison of Phusion vs Q5 DNA Polymerases for amplification of HSV1. Amplification of serially diluted HSV1 (5.23x 10^5^ to 5.23×10^2^ copies/reaction) in triplicates with (A) Phusion-LOD of 5.23×10^4^ copies and with (B) Q5-LOD of 5.23×10^3^ copies.

Q5 Polymerase showed better experimental sensitivity for the viral assay and has a higher theoretical fidelity (http://www.neb.com) than Phusion. It was thus chosen as the DNA Polymerase of choice for the viral assay.

### Optimization of annealing temperature

A temperature gradient PCR was performed with HCMV and HSV1 to find the optimum annealing temperature. At the highest annealing temperature of 71.7°C, amplification was observed at all concentrations of the viruses, including at an order of 10^2^ copies/reaction. As the annealing temperature decreased to 67.4°C in a gradient, amplification at 10^2^ copies/reaction decreased until the melt peak was not observed at all. Simultaneously, at lower temperatures, the occurrence and concentration of primer dimers increased. Thus, 71.7°C, approximated to 72°C, was taken as the optimized annealing temperature for the assay. Figure 6 shows melt curves for HCMV at the gradient temperatures. Similar results were observed for HSV1 (Figure 7).

**Figure 6:**
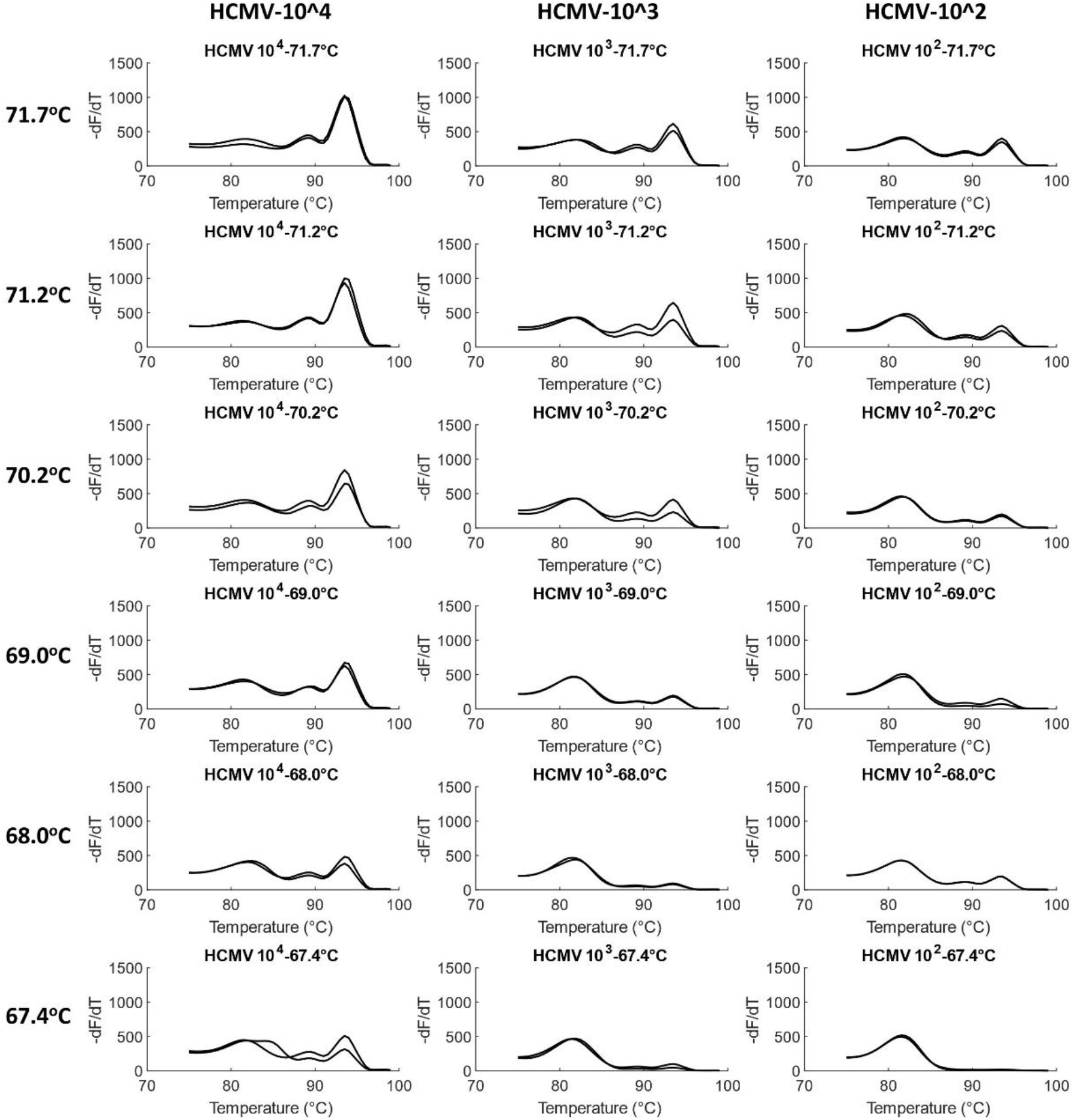
Optimization of annealing temperature in HCMV. Temperature gradient experiments were performed for HCMV in duplicates from 72°C to 67°C (rounded off) for a 10x dilution series ranging from 1.53×10^4^ to 1.53×10^2^ genome copies/reaction. Product concentration decreases with decrease in temperature, including for the lowest concentration of 1.53×10^2^ copies/reaction. ∼72°C was thus chosen as the optimum annealing temperature.

**Figure 7:**
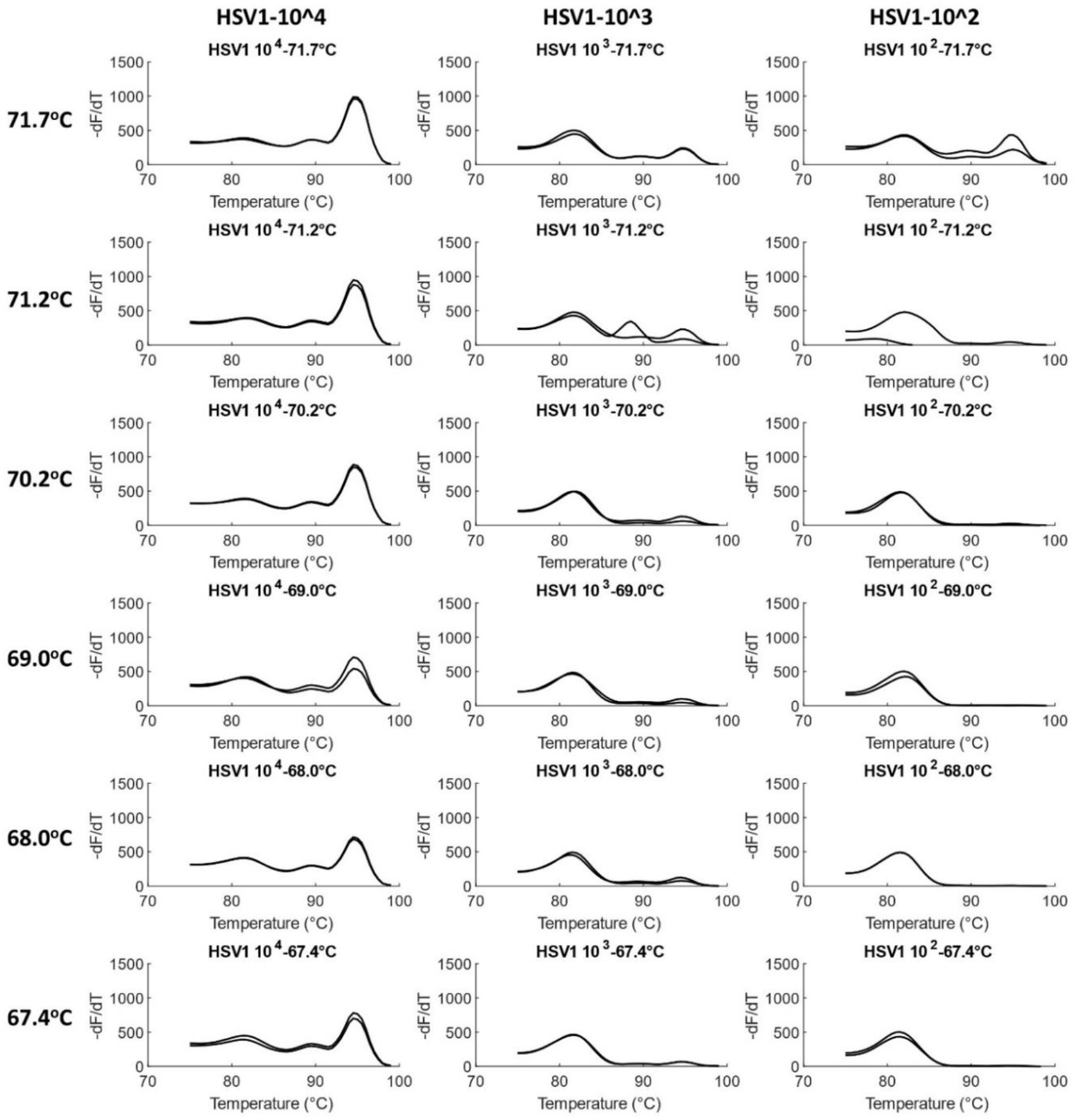
Optimization of annealing temperature in HSV1. Temperature gradient experiments were performed for HSV1 in duplicates from 72°C to 67°C (rounded off) for a 10x dilution series ranging from 5.23×10^4^ to 5.23×10^2^ genome copies/reaction. As in the case of HCMV, product concentration decreases with decrease in temperature. ∼72°C was thus chosen as the optimum annealing temperature.

Another observation was the improvement of HCMV LOD at 70°C from 1.53×10^3^ copies/reaction in the previous experiment for DNA Polymerase optimization (Figure 4) to 1.53×10^2^ copies/reaction in the present section. The current experiment was performed in Bio-Rad CFX96 qPCR instrument that was found to have a higher temperature ramp rate during thermocycling. The previous experiment, on the other hand, was performed on ThermoFisher Scientific QuantStudio where a lower ramp rate had been utilized. Therefore, in future experiments, a higher ramp rate was employed in the QuantStudio instrument, which improved the HCMV LOD to 1.53×10^2^ or 153 copies.

### Optimization of primer concentration

Initial standard curve experiments for both HCMV and HSV1 showed the limit of quantification at least an order higher (poorer) than the limit of detection observed above**-**for HCMV, LOQ was 8.45×10^3^ copies/reaction in standard curve while LOD from optimization experiments (Figure 6) was 1.53×10^2^ and for HSV1, LOQ was 5.23×10^4^ and LOD, 5.23×10^2^ (Figure 7). At concentrations lower than the LOQ, the standard curve showed a long horizontal plateau phase, but the fluorescence did not decrease to zero, as it is expected to in ideal chemistries. This occurs since the primer, p19p4, forms primer dimers even in no template controls (Figure 3A). At template concentrations below LOQ, free and unutilized primers could be forming dimers, resulting in the fluorescence observed in standard curves below LOQ. Primer dimer bands in gel electrophoresis were also observed at a template concentration of 10^5^ copies/reaction of virus for 0.5µM of each primer indicating that even at this high template concentration, either some amount of primers was left unutilized or primer dimer formation was completing with product amplification.

Therefore, an experiment was designed to test if decreasing primer concentration resulted in decreased primer dimer formation such that the product concentration was unaffected. This would also improve the LOQ of the assay. It was observed that at certain lower concentrations of primers, even though primer dimer formation decreased, so did the amount of product formation (Figure 8). The standard curve LOQ worsened by 2 orders of magnitude at 0.15µM of each primer as compared to 0.5µM (not shown). Thus 0.5µM was considered the optimized concentration for both forward and reverse primers for the assay chemistry.

**Figure 8:**
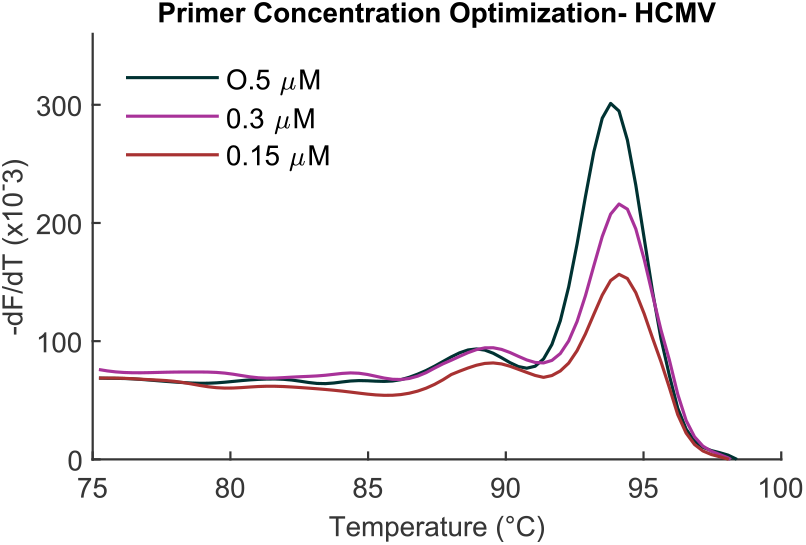
Optimizing primer concentration. Decreasing primer concentrations from 0.5 to 0.15µM decreased the amount of product formed.

The optimized probe-free viral assay thus has 0.5µM/reaction of p19p4 primer and uses Q5 DNA Polymerase. The optimum thermocycling occurs for 30 seconds annealing at 72°C and 30 seconds of extension. The ramp rates were 3°C/s for initial denaturation and cycle denaturation stages, 2.5°C/s annealing stage, 3°C/s for extension stage and final extension after the cycling, and 0.2°C/s for the final melting stage. The total run time of the assay is 1 hour 38 minutes, with an additional 10 minutes for initial ramping and final cooling.

A limitation of this optimized assay is the formation of primer dimers even in NTC (Figure 3A), which affects its analytical characteristics, as discussed below.

### Testing the optimized assay with mock samples

The optimized assay was then validated with mock clinical samples to monitor the effect of common clinical inhibitors on its sensitivity. Although the extracted viral DNA was obtained pre-purified from the vendor, blood-extracted human DNA was added to the reaction to simulate a mock clinical sample; an actual viral extraction from blood would result in a similar presence of human DNA and PCR inhibitors. 10X dilution series of HCMV DNA was created from 1.53x 10^5^ to 1.53x 10^2^ copies/reaction and ∼1.39x 10^5^ copies/reaction of extracted human DNA were added to each dilution. Each dilution was amplified in duplicates.

The addition of extracted human DNA to the assay did not inhibit the reaction chemistry. The melt curves and gel show similar sensitivity of the HCMV assay as was observed in previous sections (Figure 9). Specificity of the HCMV assay against human DNA is also underscored by this experiment.

**Figure 9:**
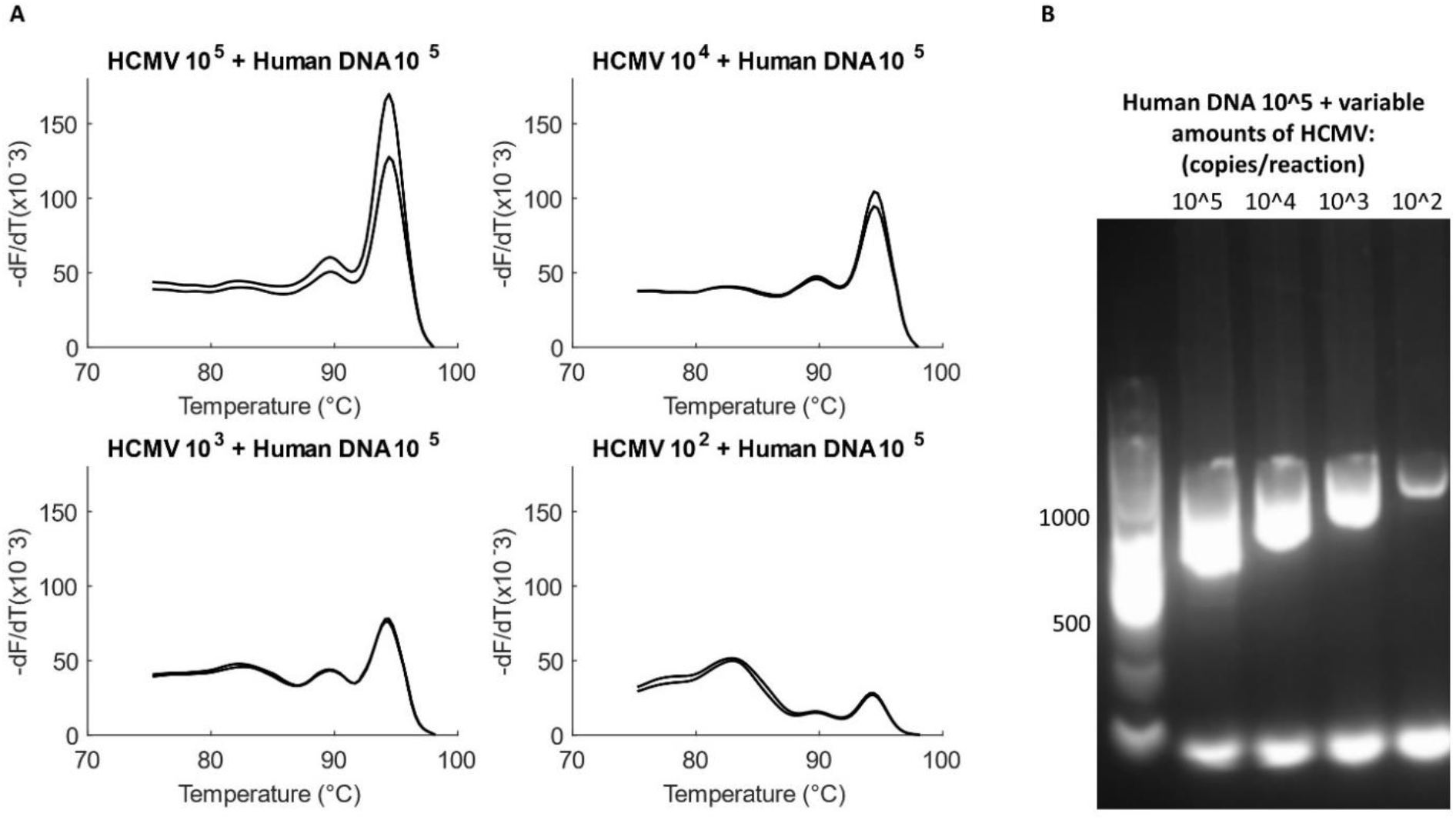
Testing the viral assay with mock samples. (A) Serial dilutions of HCMV (1.53×10^5^ to 1.53×10^2^ copies/reaction) added with the same amount of human DNA (1.39×10^5^ copies/reaction), tested in duplicates. HCMV product formation is uninhibited even at the lowest template concentration of 1.53×10^2^ copies/reaction. (B) Gel does not show extra bands for amplification of human DNA. Products and primer dimers are both observed at all the tested concentrations of the virus.

### Analytical characterization of the viral assay

Standard curves were created for the three viruses for the analytical characterization of the assay. The LOQ for HCMV is 8.45×10^3^ genome copies/reaction while it is 5.23×10^4^ copies/reaction for HSV1 and 3.45×10^4^ copies/reaction for HSV2. Figure 10 shows the standard curves for the three viruses, with the linear dynamic range marked with a linear fitted line and a plateau phase being observed at lower template concentrations. The linear dynamic range of HSV2 is poorer than the other two viruses since higher concentration of the virus showed inconsistent PCR generally attributed to the presence of inhibitors in the sample. The assay may need to be reoptimized to increase PCR performance for HSV2 or a new HSV2 sample can be ordered to check the validity of the assay. Table 3 shows the characteristics of the assay, including R^2^ values, efficiency and slope in addition to linear dynamic range and LOQ. The efficiency of the assay ranges from 85% for HSV1, 77% for HCMV to 58% for HSV2. The low efficiency could result due to utilization of longer amplicon chemistry or the formation of primer dimers. Our primer design targeted longer amplicons to preferentially detect viable over dead organisms (30, 31).

**Table 3:**
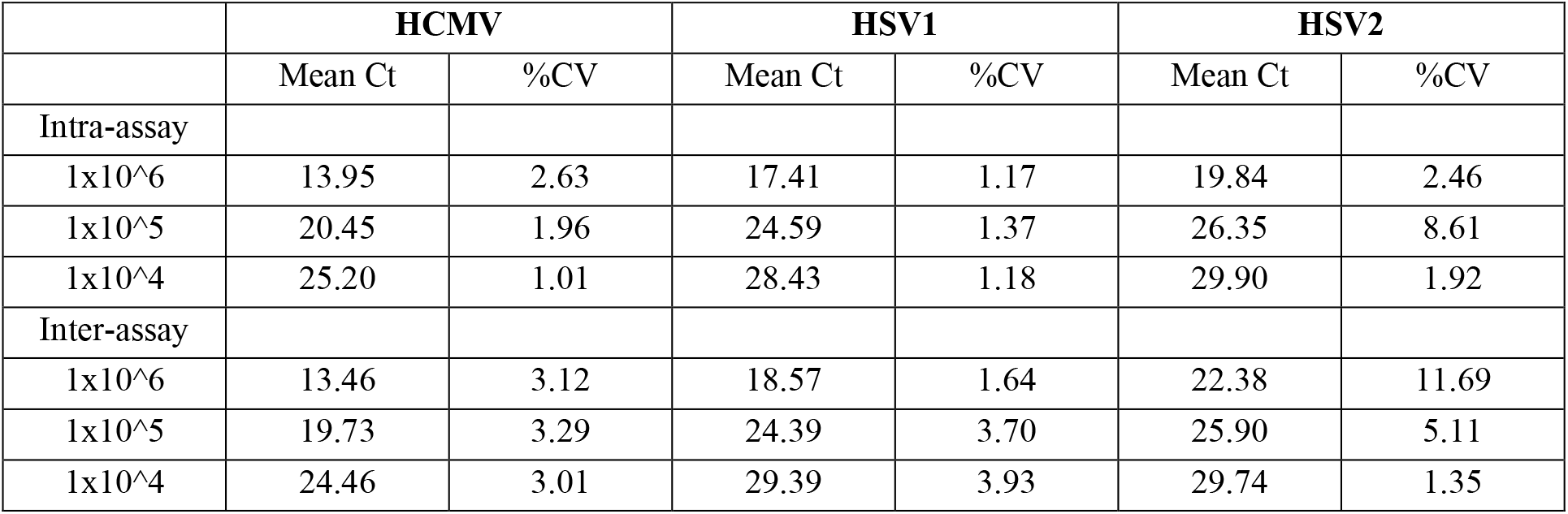
Inter and intra-assay variation for the three viruses

**Figure 10:**
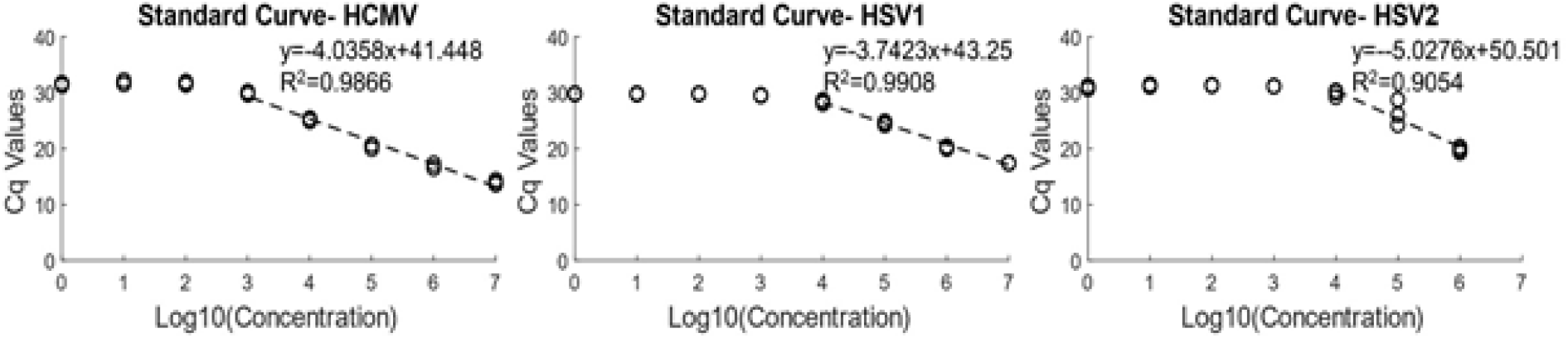
Standard curves for HCMV, HSV1 and HSV2. The quantification region (LOQ) is marked with a dashed line while the plateau region at lower concentrations is due to primer dimers. The serial dilution concentrations are as follows: HCMV-8.45×10^7^ to 8.45×10° copies/reaction, HSV1-5.23×10^7^ to 5.23×10° copies/reaction and HSV2-3.45×10^6^ to 3.45×10° copies/reaction.

Intra and inter-assay variation was found with three repeat standard curve experiments. For HCMV and HSV2, each experiment was with triplicates while for HSV1, one experiment (for determining intra-assay variation) was in triplicates while the other two were duplicates. The inter- and intra-assay coefficient of variation for HCMV and HSV1 were low, ranging from 1.01 to 2.82% (intra assay) and from 3.01 to 5.98% (inter-assay). The HSV2 experiments had higher intra- and inter-assay variation (Table 4).

## Discussion

HCMV, HSV1 and HSV2 are the leading causes of congenital viral infections. These infections can range in severity from being asymptomatic to life-threatening, and may cause devastating long-term neurodevelopmental impairments in survivors (32, 33). Because these infections may be asymptomatic after birth or present with non-specific clinical findings, a failure to correctly diagnose the condition may increase the infant’s risk for developing late onset morbidity and/or mortality (1, 12, 34). There is a need for a rapid, inexpensive, specific and multiplexed diagnostic technology for these viruses that can enable early screening and correct diagnosis for better patient management. The gold standard for herpesviral detection is viral culture (35, 36), which has a higher turn-around time, higher costs and poorer sensitivity (36). These attributes make culture methodologies unsuitable as a tool for rapid and multiplexed screening. Probe-free PCR along with high resolution melt offers a rapid, inexpensive and specific detection technology amenable to multiplexing that may alleviate some of the problems of current detection as highlighted above.

This work’s aim was to develop and optimize a probe-free viral assay for HCMV, HSV1 and HSV2. While a few universal primers for these viruses are available in literature (29, 37), these are either non-specific against human DNA amplification or had high degeneracy/mismatches with the template that would affect the specificity and the stability of the reaction (38). Custom primers were therefore designed for the assay and optimization performed. The optimized assay was found to portray specificity for each of the three viruses with distinct melt curve signatures for each (Figure 3C). Specificity against human DNA amplification was also observed. The optimized assay has a total turn-around time of <4 hours (2 hours for DNA extraction and sample preparation and <2 hours for thermocycling and melt imaging). The LOD of the assay is 1.53×10^2^ copies/reaction or 10,200 genome copies/ml for HCMV and 5.23×10^2^ copies/reaction or 34,867 genome copies/ml for HSV1 while the LOQ from standard curves was found to be in the orders of 10^3^ copies/reaction for HCMV and 10^4^ for HSV1 (Figure 10). The current LOD values lie within the clinical range of viral load observed in neonates that develop sequelae (39, 40), and the detection limit may improve further on translation of the assay to a digital format.

Thus, a probe-free viral assay was developed that utilizes high resolution melt signatures for specific detection of each pathogenic species, removing the need for sequence-specific probes or nested PCR, and thus increasing the capacity for high throughput detection. The speed and affordability of the assay as compared to viral cultures would allow universal viral screening to be adopted, which is particularly advantageous for cCMV and maternal HSV infections (1, 18).

Notwithstanding the advantages of the assay in clinical neonatal diagnostics, more work needs to be done in addressing the limitations of the assay. The custom primer pair forms primer dimers in NTC (Figure 3A) that seems to be affecting the assay efficiency, LOQ and LOD and linear dynamic range. The limitations of poor efficiency could potentially be solved by translating the qPCR assay to a digital PCR (dPCR) platform, since dPCR is an endpoint assay that tolerates longer cycling without loss of quantitative power (41). The assay also shows better performance for HSV1 and CMV than for HSV2, as observed by lower efficiency and poorer linear dynamic range for HSV2. It thus requires more optimization in particular for HSV2, which might improve its analytical characteristics. Nonetheless, the assay developed herein could enable specific diagnosis and faster clinical decision-making to reduce morbidity from late onset conditions by timely interventions, thus reducing overall healthcare costs.

A future aim for the viral assay would be its multiplexing with our lab’s bacterial and fungal sepsis assays to obtain a single multiplexed diagnostic panel for different pathogenic etiologies in cases of clinically ambiguous symptoms. The probe-free chemistry of the assay would allow easier multiplexing where the species level specificity would be attained through melt signatures. Translation of the multiplexed assay to our lab’s digital molecular technology (digital PCR or dPCR) would further increase its sensitivity to single genome level along with broad based detection, employing the lab’s machine learning algorithm to “learn” unique melt signatures of a multitude of amplicons. This would also result in the usage of a smaller sample volume than traditional PCR.

## Data Availability

Available upon request

## Acknowledgements

S.I.F. and S.M.L. envisioned the study, A.G. designed and implemented the study, A.G. and S.I.F. analyzed data, A.G. wrote the paper, all authors edited the paper. Funding was provided by NIH/NICHD funding 1R01HD099250-01A1.

## References

1. Cannon MJ, Griffiths PD, Aston V, Rawlinson WD. 2014. Universal newborn screening for congenital CMV infection: What is the evidence of potential benefit? Rev Med Virol 24:291–307.

2. A. Ross S, Novak Z, Pati S, B. Boppana S. 2012. Overview of the Diagnosis of Cytomegalovirus Infection. Infect Disord - Drug Targets 11:466–474.

3. Pass RF. 1985. Epidemiology and Transmission of Cytomegalovirus. J Infect Dis 152:243–248.

4. Davis NL, King CC, Kourtis AP. 2017. Cytomegalovirus infection in pregnancy. Birth Defects Res 109:336–346.

5. Dimech W, Cabuang LM, Grunert HP, Lindig V, James V, Senechal B, Vincini GA, Zeichhardt H. 2018. Results of cytomegalovirus DNA viral loads expressed in copies per millilitre and international units per millilitre are equivalent. J Virol Methods 252:15–23.

6. Kenneson A, Cannon MJ. 2007. Review and meta-analysis of the epidemiology of congenital cytomegalovirus (CMV) infection. Rev Med Virol 17:253–76.

7. Lanari M, Lazzarotto T, Venturi V, Papa I, Gabrielli L, Guerra B, Landini MP, Faldella G. 2006. Neonatal cytomegalovirus blood load and risk of sequelae in symptomatic and asymptomatic congenitally infected newborns. Pediatrics 117:e76–e83.

8. Manicklal S, Emery VC, Lazzarotto T, Boppana SB, Gupta RK. 2013. The “Silent” global burden of congenital cytomegalovirus. Clin Microbiol Rev 26:86–102.

9. Cannon MJ, Schmid DS, Hyde TB. 2010. Review of cytomegalovirus seroprevalence and demographic characteristics associated with infection. Rev Med Virol 20:202–213.

10. Staras SAS, Dollard SC, Radford KW, Flanders WD, Pass RF, Cannon MJ. 2006. eroprevalence of Cytomegalovirus Infection in the United States, 1988-1994. Clin Infect Dis 43:1143–1151.

11. Colugnati FAB, Staras SAS, Dollard SC, Cannon MJ. 2007. Incidence of cytomegalovirus infection among the general population and pregnant women in the United States. BMC Infect Dis 7:71.

12. Nigro G. 2009. Maternal–fetal cytomegalovirus infection: From diagnosis to therapy. J Matern Neonatal Med 22:169–174.

13. Swanson EC, Schleiss MR. 2013. Congenital Cytomegalovirus Infection. New Prospects for Prevention and Therapy. Pediatr Clin North Am 60:335–49.

14. Wald A, Corey L. 2007. HSV: Persistence in the population: Epidemiology, transmission, p. 656–672. In Arvin A, Campadelli-Fiume G, Mocarski E, Moore PS, Roizman B, Whitley R, Yamanishi K (ed), Human Herpesviruses: Biology, Therapy, and Immunoprophylaxis. Cambridge University Press, Cambridge.

15. Fatahzadeh M, Schwartz RA. 2007. Human herpes simplex virus infections: Epidemiology, pathogenesis, symptomatology, diagnosis, and management. J Am Acad Dermatol 57:737–63.

16. Whitley RJ, Roizman B. 2001. Herpes simplex virus infections. Lancet 357:1513–1518.

17. Whitley RJ, Nahmias AJ, Visintine AM, Fleming CL, Alford CA, Yeager A, Arvin A, Haynes R, Hilty M, Luby J. 1980. The Natural History of Herpes Simplex Virus Infection of Mother and Newborn. Pediatrics 66:489–4194.

18. Pinninti SG, Kimberlin DW. 2014. Preventing herpes simplex virus in the newborn. Clin Perinatol 41:945–955.

19. Kimberlin DW. 2007. Herpes Simplex Virus Infections of the Newborn. Semin Perinatol 31:19–25.

20. James SH, Sheffield JS, Kimberlin DW. 2014. Mother-to-Child Transmission of Herpes Simplex Virus. J Pediatric Infect Dis Soc 3:S19–S23.

21. James SH, Kimberlin DW. 2015. Neonatal herpes simplex virus infection: Epidemiology and treatment. Clin Perinatol 42:47–59.

22. Corey L, Wald A. 2009. Maternal and neonatal herpes simplex virus infections. N Engl J Med 361:1376–1385.

23. Lin GL, McGinley JP, Drysdale SB, Pollard AJ. 2018. Epidemiology and Immune Pathogenesis of Viral Sepsis. Front Immunol 9:2147.

24. Sinha M, Jupe J, Mack H, Coleman TP, Lawrence SM, Fraley SI. 2018. Emerging technologies for molecular diagnosis of sepsis. Clin Microbiol Rev. American Society for Microbiology.

25. Hodinka RL, Kaiser L. 2013. Is the era of viral culture over in the clinical microbiology laboratory? J Clin Microbiol 31:e00089–17.

26. Razonable RR, Inoue N, Pinninti SG, Boppana SB, Lazzarotto T, Gabrielli L, Simonazzi G, Pellett PE, Schmid DS. 2020. Clinical Diagnostic Testing for Human Cytomegalovirus Infections. J Infect Dis 221:S74–S85.

27. Johnson G, Nelson S, Petric M, Tellier R. 2000. Comprehensive PCR-based assay for detection and species identification of human herpesviruses. J Clin Microbiol 38:3274– 3279.

28. Mocarski Jr. ES. 2007. Comparative analysis of herpesvirus-common proteins, p44–58. In Arvin A, Campadelli-Fiume G, Mocarski E, Moore PS, Roizman B, Whitley R, Yamanishi K (ed), Human Herpesviruses: Biology, Therapy, and Immunoprophylaxis. Cambridge University Press, Cambridge.

29. Rozenberg F, Lebon P. 1991. Amplification and Characterization of Herpesvirus DNA in Cerebrospinal Fluid from Patients with Acute Encephalitis. J Clin Microbiol 29:2412–2417.

30. Mccarty SC, Atlas RM. 1993. Effect of Amplicon Size on PCR Detection of Bacteria Exposed to Chlorine. PCR Methods Appl 3:181–185.

31. Immanuel T, Taylor R, Keeling S, Brosnahan C, Alexander B. 2020. Discrimination between viable and dead Xanthomonas fragariae in strawberry using viability PCR. J Phytopathol 168:363–373.

32. Van Lint AL, Knipe DM. 2019. Herpesviruses, p. 565–579. In Schmidt T (ed) Encyclopedia of Microbiology, 4th ed. Elsevier, Cambridge, MA.

33. Cantey JB. 2018. Pathogenesis of Congenital Infections, p67–73. In Cantey JB (ed) Neonatal Infections Pathophysiology, Diagnosis, and Management. Springer, New York, NY.

34. Shane AL, Sánchez PJ, Stoll BJ. 2017. Neonatal sepsis. Lancet 390:1770–80.

35. Harris JB, Holmes AP. 2017. Neonatal herpes simplex viral infections and acyclovir: An update. J Pediatr Pharmacol Ther 22:88–93.

36. Ross SA, Ahmed A, Palmer AL, Michaels MG, Sánchez PJ, Bernstein DI, Tolan RW, Novak Z, Chowdhury N, Fowler KB, Boppana SB, National Institute on Deafness. 2014. Detection of congenital cytomegalovirus infection by real-time polymerase chain reaction analysis of saliva or urine specimens. J Infect Dis 210:1415–1418.

37. Sankuntaw N, Sukprasert S, Engchanil C, Kaewkes W, Chantratita W, Pairoj V, Lulitanond V. 2011. Single tube multiplex real-time PCR for the rapid detection of herpesvirus infections of the central nervous system. Mol Cell Probes 25:114–120.

38. Stadhouders R, Pas SD, Anber J, Voermans J, Mes THM, Schutten M. 2010. The effect of primer-template mismatches on the detection and quantification of nucleic acids using the 5′ nuclease assay. J Mol Diagnostics 12:109–117.

39. Forner G, Abate D, Mengoli C, Palù G, Gussetti N. 2015. High cytomegalovirus (CMV) DNAemia predicts CMV sequelae in asymptomatic congenitally infected newborns born to women with primary infection during pregnancy. J Infect Dis 212:67–71.

40. Melvin AJ, Mohan KM, Schiffer JT, Drolette LM, Magaret A, Corey L, Wald A. 2015. Plasma and cerebrospinal fluid herpes simplex virus levels at diagnosis and outcome of neonatal infection. J Pediatr 166:827–833.

41. Sinha M, Mack H, Coleman TP, Fraley SI. 2018. A High-Resolution Digital DNA Melting Platform for Robust Sequence Profiling and Enhanced Genotype Discrimination. SLAS Technol 23:580–591.

